# Classification and Visualisation of Normal and Abnormal Radiographs; a comparison between Eleven Convolutional Neural Network Architectures

**DOI:** 10.1101/2021.06.16.21259014

**Authors:** Ananda Ananda, Kwun Ho Ngan, Cefa Karabağ, Aram Ter-Sarkisov, Eduardo Alonso, Constantino Carlos Reyes-Aldasoro

**Affiliations:** giCentre, Department of Computer Science, School of Mathematics, Computer Science and Engineering, City, University of London, London EC1V 0HB, UK; CitAI Research Centre, Department of Computer Science, School of Mathematics, Computer Science and Engineering, City, University of London, London EC1V 0HB, UK

**Keywords:** Wrist Fractures, Radiographic Images, Classification, Convolutional Neural Networks, Class Activation Mapping

## Abstract

This paper investigates the classification of radiographic images with eleven convolutional neural network (CNN) architectures (*GoogleNet, VGG-19, AlexNet, SqueezeNet, ResNet-18, Inception-v3, ResNet-50, VGG-16, ResNet-101, DenseNet-201 and Inception-ResNet-v2*). The CNNs were used to classify a series of wrist radiographs from the Stanford Musculoskeletal Radiographs (MURA) dataset into two classes - normal and abnormal. The architectures were compared for different hyper-parameters against accuracy and Cohen’s kappa coefficient. The best two results were then explored with data augmentation. Without the use of augmentation, the best results were provided by Inception-Resnet-v2 (Mean accuracy = **0.723**, Mean kappa = **0.506**). These were significantly improved with augmentation to Inception-Resnet-v2 (Mean accuracy = **0.857**, Mean kappa = **0.703**). Finally, Class Activation Mapping was applied to interpret activation of the network against the location of an anomaly in the radiographs.

## Introduction

Fractures of the wrist and forearm are common injuries, especially among older and frail persons who may slip and extend the arm to protect themselves [1]. In some cases, the person involved may think that they have not injured themselves seriously and the fractures are ignored and left untreated [2]. These fractures can provoke impairment in the wrist movement [3]. In more serious cases, fractures can lead to complications such as ruptured tendons or long-lasting stiffness of the fingers [4] and can impact the quality of life [5].

Treatment of fractures through immobilisation and casting is an old, tried-and-tested technique. There are Egyptian records describing the re-positioning of bones, fixing with wood and covering with linen [6] and there are also records of fracture treatment in Iron Age and Roman Britain where “skilled practitioners” treated fractures and even “minimised the patient’s risk of impairment” [7]. The process of immobilisation is now routinely performed in the Accidents and Emergency (A&E) departments of hospitals under local anaesthesia and is known as *Manipulation under Anaesthesia* (MUA) [8], or closed reduction and casting. MUA interventions in many cases represent a significant proportion of the Emergency Department workload. In many hospitals, patients are initially treated with a temporary plaster cast, then return afterwards for the manipulation as a planned procedure. MUA, although simple, is not entirely free of risks. Some of the problems include bruising, tears of the skin, complications related to the local anaesthetic and there is discomfort for the patients. It should be noted that a large proportion of MUA procedures fail. It has been reported that 41% of Colles’ fractures treated with MUA required alternative treatment [9]. The alternative to MUA is open surgery, which is also known as *Open Reduction and Internal Fixation* (ORIF) [10], and can be performed with local or general anaesthesia [11,12] to manipulate the fractured bones and fixate them with metallic pins, plates or screws. The surgical procedure is more complicated and expensive than MUA. In some cases, it can also lead to serious complications especially with metallic elements that can interfere with the tendons and cut through subchondral bones [13,14]. ORIF it is more reliable as a long-term treatment.

Despite the considerable research in the area ([8,10,13,15–18]), there is no certainty into which procedure to follow for wrist fractures [19–21]. The main tool to examine wrist fractures is through diagnostic imaging, e.g., X-ray or Computed Tomography (CT). The images produced are observed by highly skilled radiologist and radiographers in search for anomalies, and based on experience, they then determine the most appropriate procedure for each case. The volume of diagnostic images has increased significantly [22], and work overload is further exacerbated by a shortage of qualified radiologists and radiographers as exposed by The Royal College of Radiologists [23]. Thus, the possibility of providing computational tools to assess radiographs of wrist fractures is attractive. Traditional analysis of wrist fractures has focused on geometric measurements that are extracted either manually [24–27] or through what is now considered traditional image processing [28]. The geometric measurements that have been of interest are, amongst others: radial shortening [29], radial length [25], volar and dorsal displacements [30], palmar tilt and radial inclination [31], ulnar variance [24], articular stepoff [26], and metaphyseal collapse ratio [27]. Non-geometric measurements such as bone density [32,33] as well as other osteoporosis-related measurements e.g., cortical thickness, internal diameter, cortical area [34] have also been considered to evaluate bone fragility.

However, in recent years, computational advances have been revolutionised by the use of machine learning and artificial intelligence (AI), especially with *deep learning architectures* [35]. Deep learning is a part of the machine learning methods where input data is provided to a model to discover or learn the representations that are required to perform a classification [36]. These models have a large number levels, far more than the input/hidden/output layers of the early configurations and thus considered *deep*. At each level, non linear modules transform the representation of the data from the input data into a more abstract representation [37].

Deep learning has had significant impact in many areas of image processing and computer vision, for instance, it provides outstanding results in difficult tasks like the classification of the ImageNet Large Scale Visual Recognition Challenge (ILSVRC) [38] and it has been reported that deep learning architectures have in some cases outperformed expert dermatologists in classification of skin cancer [39]. Deep learning has been widely applied for segmentation and classification [40–48].

Deep learning applied system versus radiologists’ interpretation on detection and localisation of distal radius fractures has been reported by [49]. Diagnostic improvements have been studied by [50] where deep learning supports the medical specialist to a better outcome to the patient care. Automated fracture detection and localisation for wrist radiographs are also feasible for further investigation [51].

Notwithstanding their merits, deep learning architectures have several well-known limitations: significant computational power is required together with large amounts of training data. There is a large number of architectures, and each of them will require a large number of parameters to be fine-tuned. Many publications will use one or two of these architectures and compare against a baseline, like human observers or a traditional image processing methodology. However, a novice user may struggle to select one particular architecture, which in turn may not necessarily be the most adequate for a certain purpose. In addition, one recurrent criticism is their *black box* nature [52–55], which implies that it is not always easy or simple to understand how the networks perform in the way they do. One method to address this *opacity* is through explainable techniques, such as activation maps [56,57] as a tool explain visually the localisation of class-specific image regions.

In this work, the classification of radiographs into 2 classes, normal and abnormal, with eleven convolutional neural network (CNN) architectures was investigated. The architectures compared were the following: (*GoogleNet, VGG-19, AlexNet, SqueezeNet, ResNet-18, Inception-v3, ResNet-50, VGG-16, ResNet-101, DenseNet-201 and Inception-ResNet-v2*). This paper extends a preliminary version of this work [58]. Here, we extended the work by applying data augmentation to the two models that provided the best results, that is, ResNet-50 and Inception-ResNet-v2. Furthermore, class activation maps were generated analysed.

The dataset used to compare the architectures was the *Stanford MURA (musculoskeletal radiographs)* radiographs [59]. This is a database that contains a large number of radiographs; 40,561 images from 14,863 studies, where each study is manually labelled by radiologists as either normal/abnormal. The radiographs cover seven anatomical regions, namely Elbow, Finger, Forearm, Hand, Humerus, Shoulder and Wrist. This paper focused mainly on the wrist images. The main contributions of this work are the following: (1) an objective comparison of the classification results of 11 architectures, this can help the selection of a particular architecture in future studies, (2) the comparison of the classification with and without data augmentation, which resulted in significantly better results, (3) The use of Class Activation Mapping to analyse the regions of interest of the radiographs.

The rest of the manuscript is organised as follows. Section 2 describes the materials, that is, the data base of radiographs, and the methods that describe the Deep Learning models that were compared and the Class Activation Mapping (CAM) to visualise the activated regions. The performance metrics of accuracy and Cohen’s kappa coefficient are described at the end of this section. Section 3 present the results of all the experiments and the effect of the different hyper-parameters. Predicted abnormality in the radiographic images will also be visualised by using class activation mapping. The manuscript finishes with a discussion of the results in section 4.

## Materials and Methods

### Materials

The data used to compare the 11 CNNs was obtained from the public dataset Musculoskeletal RAdiographs (MURA) from a competition organised by researchers from Stanford University [59]. The dataset has been manually labelled by board-certified radiologists between 2001 and 2012. The studies (*n* = 14, 656) are divided into training (*n* = 13, 457), and validation (*n* = 1, 199). Furthermore, the studies have been allocated in groups called abnormal (i.e., those radiographs that contained fractured bones, foreign bodies such as implants, wires or screws, etc.) (*n* = 5, 715) or normal (*n* = 8, 941). Representative normal cases are illustrated in Fig. 1 and abnormal cases in Fig. 2. The distribution per anatomical region is shown in Table 1. In this paper, the subset of the **wrists** was selected. The cases of normal and abnormal wrist radiographs is presented in Table 2. Notice that these were subdivided into four studies.

**Table 1.** Distribution of studies of the Stanford MURA (musculoskeletal radiographs) data set [59] for studies of the upper body.

| No. | Study | Train |  | Validation |  | Total |
| --- | --- | --- | --- | --- | --- | --- |
|  |  | Normal | Abnormal | Normal | Abnormal |  |
| 1 | Elbow | 1094 | 660 | 92 | 66 | 1912 |
| 2 | Finger | 1280 | 655 | 92 | 83 | 2110 |
| 3 | Hand | 1497 | 521 | 101 | 66 | 2185 |
| 4 | Humerus | 321 | 271 | 68 | 67 | 727 |
| 5 | Forearm | 590 | 287 | 69 | 64 | 1010 |
| 6 | Shoulder | 1364 | 1457 | 99 | 95 | 3015 |
| 7 | **Wrist** | **2134** | **1326** | **140** | **97** | **3697** |
|  | Total | 8280 | 5177 | 661 | 538 | 14656 |

**Table 2.** Details of the number of wrist radiographs. Studies 1,2,3 and 4 refer to a patient visit identifier; each patient may have visited the hospital several times. A positive label, corresponds to abnormal condition, whereas negative corresponds to a normal condition as decided by the expert.

| **Wrist-Train dataset** | **Abnormal** | **Normal** |
| --- | --- | --- |
| Study 1 | 3920 | 5282 |
| Study 2 | 64 | 425 |
| Study 3 | 3 | 45 |
| Study 4 | 0 | 13 |
| **Total** | 3987 | 5765 |
| **Total Wrist Train Images** | 9752 |  |
| **Wrist-Valid dataset** | **Abnormal** | **Normal** |
| Study 1 | 287 | 293 |
| Study 2 | 5 | 59 |
| Study 3 | 3 | 9 |
| Study 4 | 0 | 3 |
| **Total** | 295 | 364 |
| **Total Wrist Valid Images** | 659 |  |
| **Total Images of Wrist** | 10411 |  |

**Figure 1.**
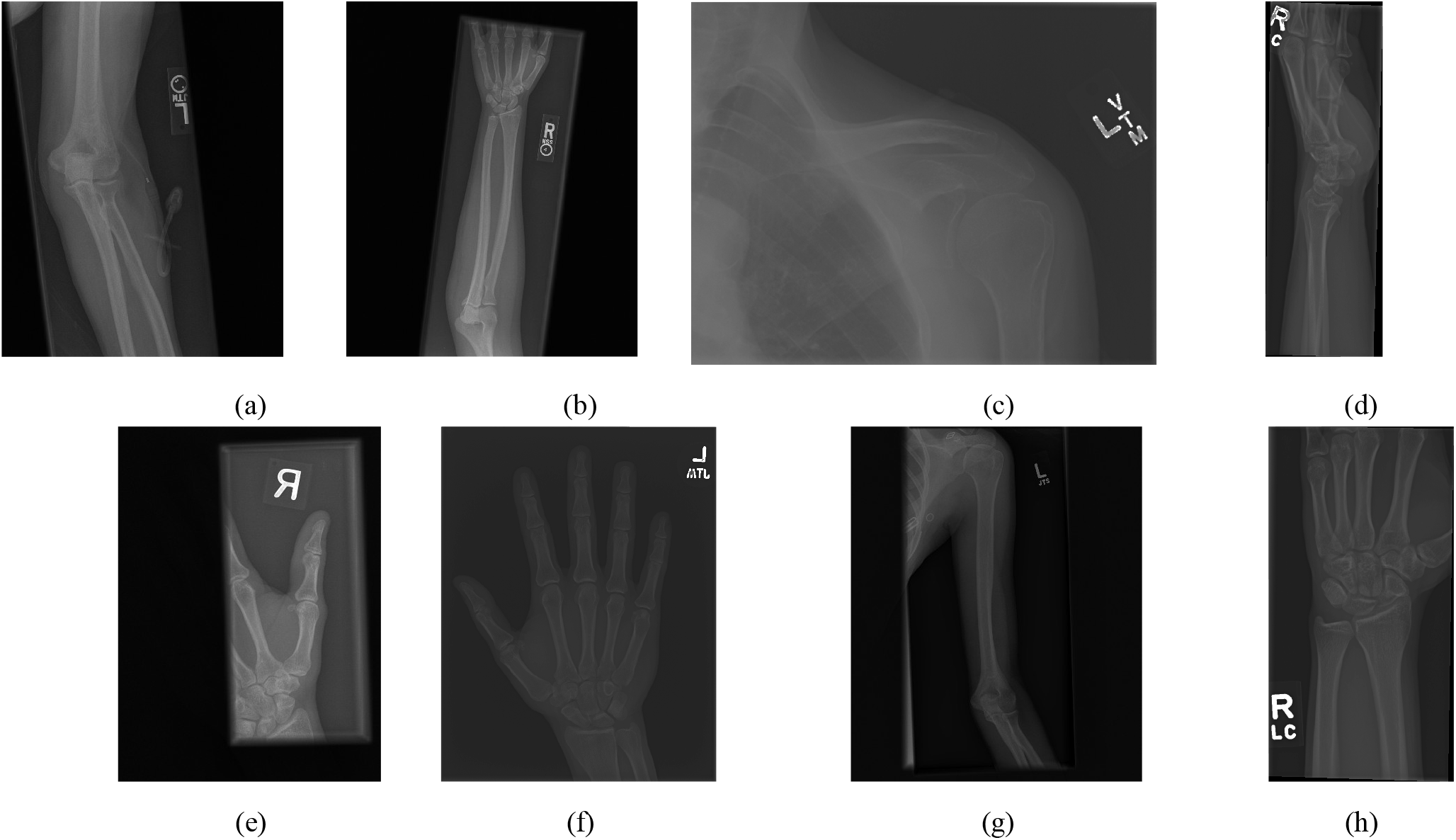
Eight examples of radiographs without abnormalities (considered negative) of the **MU**sculoskeletal **RA**diographs (MURA) dataset [59]. (a) Elbow, (b) Forearm, (c) Shoulder, (d) Wrist (lateral view), (d) Lateral view of Wrist, (e) Finger, (f) Hand, (g) Humerus, (h) Wrist. It should be noted the variability of the images in terms of dimensions, quality, contrast and the large number of labels (i.e.,R for right and L for left), which appear in various locations.

**Figure 2.**
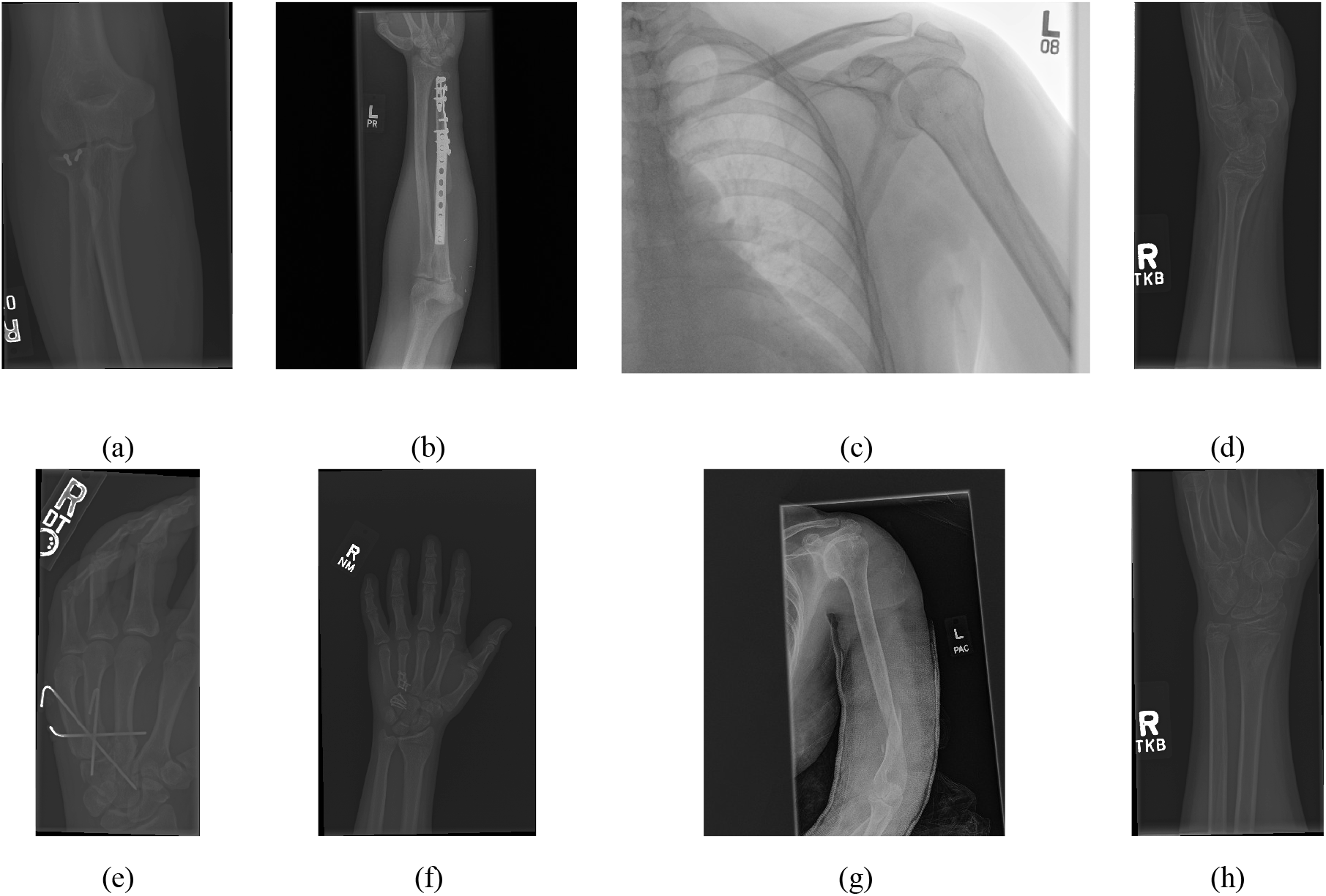
Eight examples of radiographs with abnormalities (considered positive) of the **MU**sculoskeletal **RA**diographs (MURA) dataset [59]. (a) Elbow, (b) Forearm, (c) Shoulder, (d) Wrist (lateral view), (d) Lateral view of Wrist, (e) Finger, (f) Hand, (g) Humerus, (h) Wrist. As for the cases without abnormalities, it should be noted the variability of the images and in addition the abnormalities themselves. There are cases of metallic implants some of which are smaller (a) than others (b), as well as fractures.

### Convolutional Neural Network

Convolutional Neural Networks (CNN) is a type of deep learning [35,36] models. A typical CNN classification model is composed of two key components: first, feature are extracted though a series of convolutional layers with pooling and activation functions. Some modern architectures (e.g. ResNet) will also include batch normalization and/or skip connections to mitigate the problem of vanishing gradient during model training. Next, these features input to one or more fully-connected layers to derive the final classification prediction (e.g. an estimated class probability). These class predictions are used to compute the problem-specific loss.

The input in a CNN, i.e., an image to be classified, can be transformed through the feature extraction layers to form a set of relevant features required by the network. These features can be regarded as the global descriptors of the image. In the fully-connected layers for classification, the relations of the features are learned by an iterative process of weight adjustment. A prediction probability can be deduced at the final layer with the inclusion of an activation function (e.g., softmax function). At the training stage, a loss (e.g. cross entropy loss) is computed between the prediction and the ground truth for weight adjustment during backpropagation. At the evaluation stage, the predicted class can be inferred from most probable class using an argmax function and this can be evaluated against the ground truth for classification accuracy.

A description summary of the applied models used in Table. 3 is as follows: AlexNet [60] is one of the earlier adoptions of deep learning in image classification and has won the ILSVRC 2012 competition by significantly outperformed its next runner up. It consists of 5 layers of convolutions of various sizes and 3 fully connected layers. It also applies a ReLU activation for nonlinearity. GoogleNet (Inception V1) [61] introduced the inception module formed of small size convolutions to reduce trainable parameters for better computational utilisation. Despite a deeper and wider network than AlexNet, the number of parameters for training has reduced from 60 million (Alexnet) to 4 million. VGG [62] is the runner-up in the ILSVRC2014 which was won by GoogleNet in the same year. It utilises only 3×3 convolutions in multiple layers and is deeper than AlexNet. It has a total of 138 million trainable parameters and thus can be computationally intensive during training. ResNet [63] is formed by a deep network of repetitive residual blocks. These blocks are made up of multiple convolution layers coupled with a skip connection to learn the residual based on the previous block. This allows the network to be very deep capable of 100s of network layers. Inception-v3 [64] improves the configuration of the inception module in GoogleNet from a 5×5 convolutional layer in one of the branches to two 3×3 layers reducing the number of parameters. SqueezeNet [65] introduced the fire module which consists of a layer with 1×1 convolution (i.e. squeeze layer) and a second layer with 3×3 convolution (i.e. expand layer). The number of channels into the expand layer is also reduced. This has led to a significant reduction in trainable parameters while maintaining similar accuracy to AlexNet in the ILSVRC 2012 dataset. DenseNet [66] is composed of multiple dense blocks (small convolutional layers, batch normalisation and ReLU Activation). A transition layer with batch normalisation, 1×1 convolution and average pooling is added in between the dense blocks. The blocks are each closely connected with all previous blocks by skip connections. DenseNet has demonstrated a full utilisation of residual mechanism while maintaining model compactness to achieve competitive accuracy. Inception-ResNet-v2 [67] incorporates the advantages of the Inception modules into the residual blocks of a ResNet and achieve even more accurate classification in ILSVRC 2012 dataset than either ResNet 152 or Inception-v3.

### Experiments

In this work we considered the following eleven CNN architectures to classify wrist radiographs into two categories (Normal / Abnormal): GoogleNet, VGG-19, AlexNet, SqueezeNet, ResNet-18, Inception-v3, ResNet-50, VGG-16, ResNet-101, DenseNet-201 and Inception-ResNet-v2. The details of these are presented in Table 3. The training process of the architecture was tested with different numbers of epochs (10, 20, 30), and different mini-batch sizes (16, 32, 64). The experiment pipeline is illustrated in Figure. 3. All the architectures were compared under the same conditions, without pre- or post-processing initially except resizing of the initial images to the input size for each architecture as the X-ray images presented different sizes. For instance, the images were resized to 224 × 224 for ResNet-50 and 299 × 299 for Inception-ResNet-v2. In the cases where the input was a 3-channel image, i.e., an RGB colour image, and the input image was in grayscale, this channel was replicated. The dataset was split into 90% for training and 10% for testing. The same hyper-parameters were applied as described in Table 4 and continued in Table 5.

**Table 3.** Details of convolutional neural networks (CNNs) that were used in this work.

| No. | Network | Depth | Image Input Size | Reference |
| --- | --- | --- | --- | --- |
| 1 | GoogleNet | 22 | 224-by-224 | [61] |
| 2 | VGG-19 | 19 | 224-by-224 | [62] |
| 3 | AlexNet | 8 | 227-by-227 | [60] |
| 4 | SqueezeNet | 18 | 227-by-227 | [65] |
| 5 | ResNet-18 | 18 | 224-by-224 | [63] |
| 6 | Inception-v3 | 48 | 299-by-299 | [64] |
| 7 | ResNet-50 | 50 | 224-by-224 | [63] |
| 8 | VGG-16 | 16 | 224-by-224 | [62] |
| 9 | ResNet-101 | 101 | 224-by-224 | [63] |
| 10 | DenseNet-201 | 201 | 224-by-224 | [66] |
| 11 | Inception-ResNet-v2 | 164 | 299-by-299 | [67] |

**Table 4.** Summary of convolutional neural networks (CNNs) hyper-parameters for this work.

|  |  |  |  |  |  |
| --- | --- | --- | --- | --- | --- |
| 1 | GoogleNet | Optimiser | SGDM | ADAM | RMSprop |
|  |  | Epoch | 30 | 30 | 30 |
|  |  | Mini batch size | 64 | 64 | 64 |
|  |  | Init. Learn. R. | 0.01 | 0.001 | 0.001 |
|  |  | Momentum | 0.9000 | - | - |
|  |  | L2 Reg. | 0.0001 | 0.0001 | 0.0001 |
| 2 | VGG-19 | Optimiser | SGDM | ADAM | RMSprop |
|  |  | Epoch | 30 | 30 | 30 |
|  |  | Mini batch size | 64 | 64 | 64 |
|  |  | Init. Learn. R. | 0.001 | 0.001 | 0.001 |
|  |  | Momentum | 0.9000 | - | - |
|  |  | L2 Reg. | 0.0001 | 0.0001 | 0.0001 |
| 3 | AlexNet | Optimiser | SGDM | ADAM | RMSprop |
|  |  | Epoch | 50 | 50 | 50 |
|  |  | Mini batch size | 128 | 128 | 128 |
|  |  | Init. Learn. R. | 0.001 | 0.001 | 0.001 |
|  |  | Momentum | 0.9000 | - | - |
|  |  | L2 Reg. | 0.0001 | 0.0001 | 0.0001 |
| 4 | SqueezeNet | Optimiser | SGDM | ADAM | RMSprop |
|  |  | Epoch | 30 | 30 | 30 |
|  |  | Mini batch size | 64 | 64 | 64 |
|  |  | Init. Learn. R. | 0.001 | 0.0001 | 0.0001 |
|  |  | Momentum | 0.9000 | - | - |
|  |  | L2 Reg. | 0.0001 | 0.0001 | 0.0001 |
| 5 | ResNet-18 | Optimiser | SGDM | ADAM | RMSprop |
|  |  | Epoch | 30 | 30 | 30 |
|  |  | Mini batch size | 64 | 64 | 64 |
|  |  | Init. Learn. R. | 0.001 | 0.0001 | 0.0001 |
|  |  | Momentum | 0.9000 | - | - |
|  |  | L2 Reg. | 0.0001 | 0.0001 | 0.0001 |
| 6 | Inception-v3 | Optimiser | SGDM | ADAM | RMSprop |
|  |  | Epoch | 10 | 10 | 10 |
|  |  | Mini batch size | 64 | 64 | 64 |
|  |  | Init. Learn. R. | 0.001 | 0.0001 | 0.0001 |
|  |  | Momentum | 0.9000 | - | - |
|  |  | L2 Reg. | 0.0001 | 0.0001 | 0.0001 |
| 7 | ResNet-50 | Optimiser | SGDM | ADAM | RMSprop |
|  |  | Epoch | 30 | 30 | 30 |
|  |  | Mini batch size | 64 | 64 | 64 |
|  |  | Init. Learn. R. | 0.001 | 0.0001 | 0.0001 |
|  |  | Momentum | 0.9000 | - | - |
|  |  | L2 Reg. | 0.0001 | 0.0001 | 0.0001 |

**Table 5.**
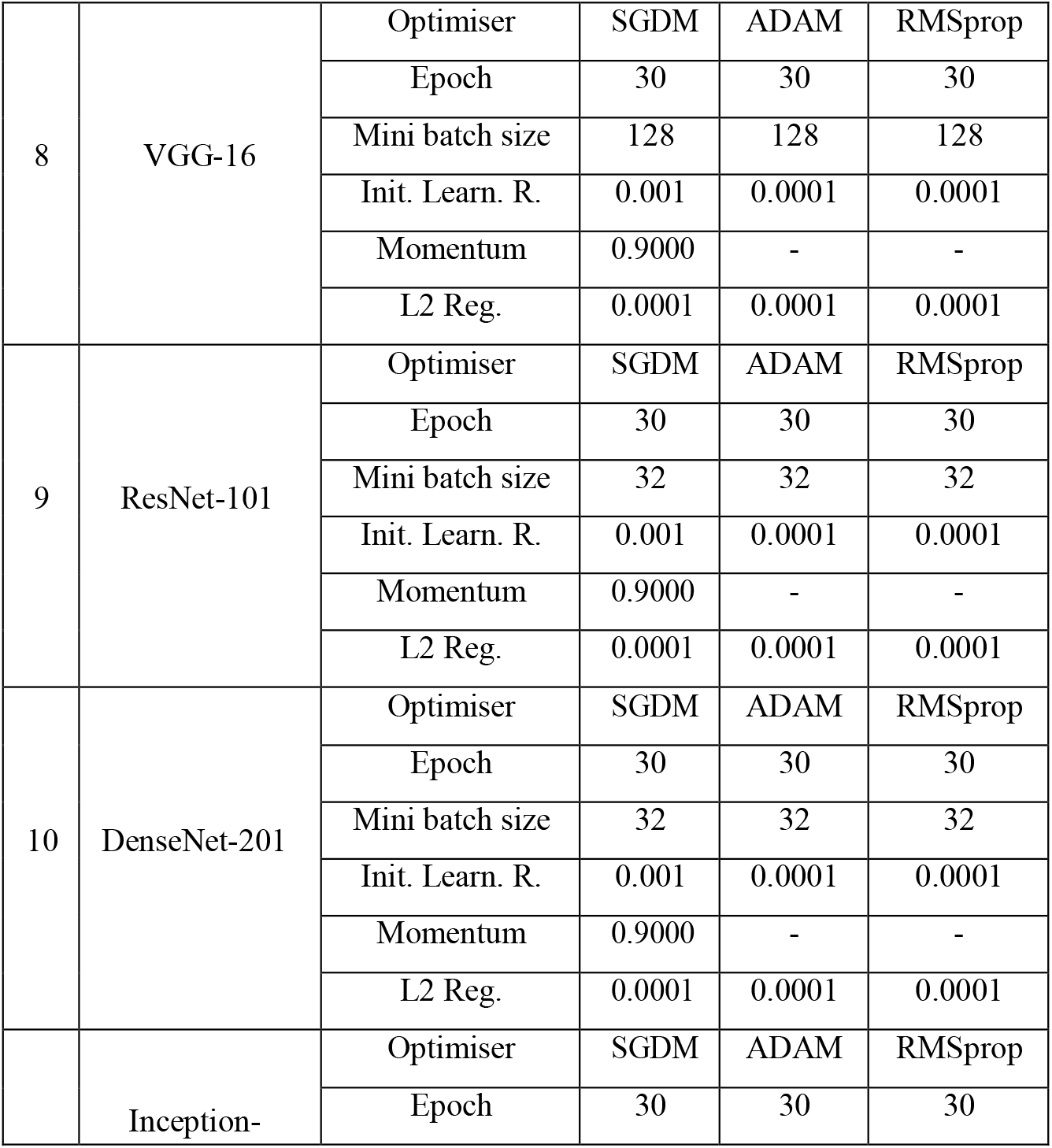
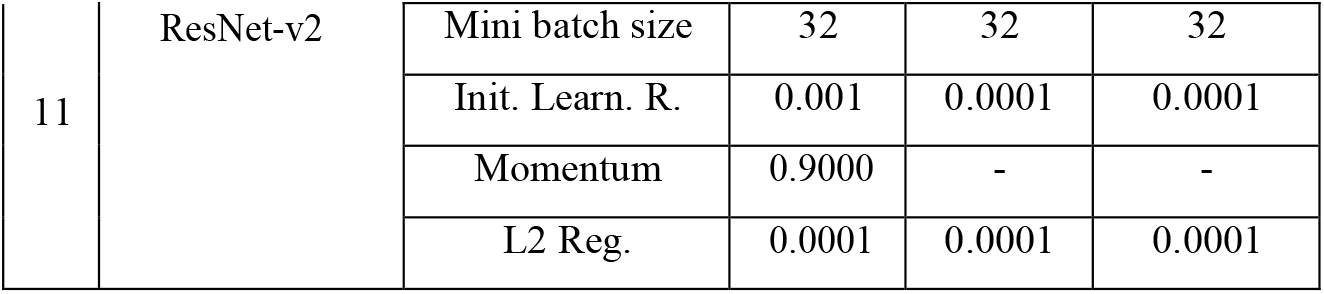
Summary of convolutional neural networks (CNNs) hyper-parameters for this work (continuation).

**Figure 3.**
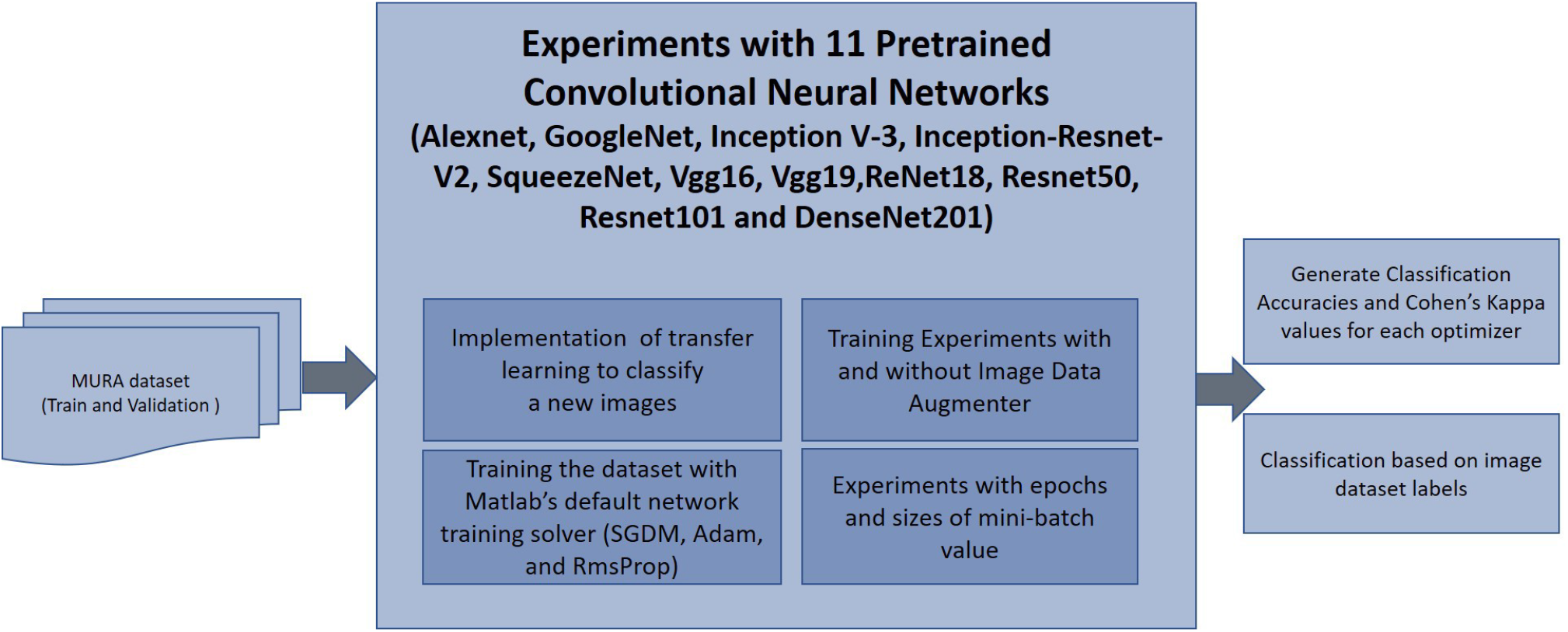
Block diagram which illustrates the classification of the wrist radiographs with 11 different Convolutional Neural Network (CNN) architectures. 9752 images from **MU**sculoskeletal **RA**diographs (MURA) Wrist dataset were used for training CNN architectures and 659 images were used for validation. Two different metrics, Accuracy (*Ac*) and Cohen’s kappa (*κ*) were computed to assess the performance of 11 pre-trained CNNs. Image data augmentation was used during training and different number of epochs and mini batch sizes were tested.

Then, for the two architectures which provided the highest accuracy and Cohen’s kappa coefficient (ResNet-50 and InceptionResnet-v2) several modifications were applied regarding, specifically, the use of data augmentation and CNN’s training options. The classification with and without augmentation was done to assess the impact that augmentation can have in the results. In addition, visualisation of the network activations with Class Activation Mapping was explored.

### Further processing with data augmentation

For the two best performing architectures, the effect of data augmentation was also be evaluated. The following augmentations have been performed to each of the training images: (1) rotations of (−5 to 5°), (2) vertical and horizontal reflections, (3) shear deformations of (−0.05 to 0.05°) in horizontal and vertical directions, and (4) Contrast-limited adaptive histogram equalisation (CLAHE) [68]. Translations were not applied as the training images were captured with a good range of translational shift.

### Class Activation Mapping

Class Activation Mapping (CAM) [56] provides a visualisation of the most significant activation mapping for a targeted class. It provides an indication of what exactly the network is focusing its attention on. Similar to the schematics in Figure 4, the class activation map is generated at the output of the last convolutional layer. In this work, this is represented with a rainbow/jet colour map where the intensity spectrum ranges from blue (lowest activation), green and red (highest activation).

**Figure 4.**
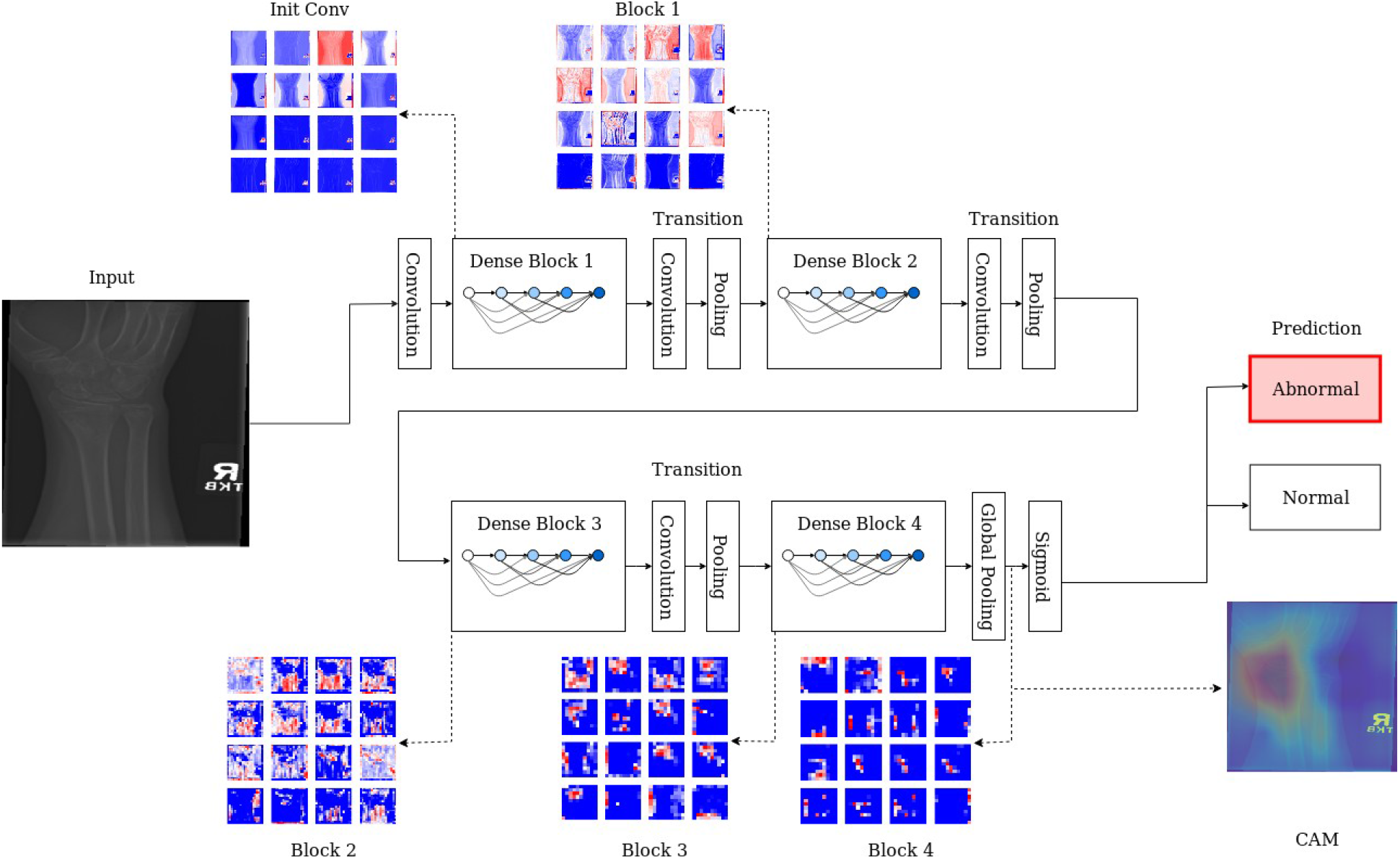
Schematic illustration of the X-ray classification process and class activation mapping through layer-wise activation maps across different dense blocks. At each level, a series of feature maps are generated, the resolution decreases progress through the blocks. Colours indicate the range of activation: blue corresponds to low activation, red for highly activated features. The final output, visualised here using Class Activation Mapping, which highlights the area(s) where abnormalities can be located.

For the two best performing models, the CAM representations were generated at layer “activation_49_relu” for ResNet-50 and “conv_7_bac” for Inception-ResNet-v2 respectively. The CAM maps were up-scaled to the input resolution and overlaid on top of the original radiography for the location of the abnormalities.

### Performance Metrics

Accuracy (*Ac*) was calculated as the proportion of correct predictions among the total number of cases examined, that is:

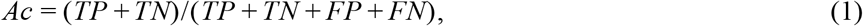

where *TP* and *TN* correspond to positive and negative classes correctly predicted and *FP* and *FN* correspond to false predictions. Cohen’s kappa (*κ*) was also calculated as it is the metric used to rank the MURA challenge [59,69] and it is considered more robust as it takes into account the possibilities of random agreements. Cohen’s kappa *κ* was calculated in the following way. With

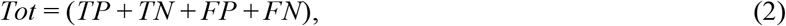

being the total number of events, the probability of a yes or *TP* is

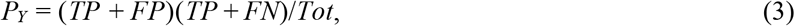

the probability of a no, or *TN* is

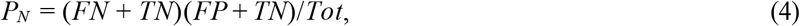

and the probability of random agreement *PR* = *PY* + *PN*, then

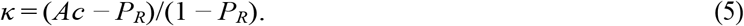

### Implementation Details

Experiments were conducted in Matlab R2018b IDE completed with Deep Learning Toolbox, Image Processing Toolbox and Parallel Computing Toolbox. These experiments were conducted using a workstation with a processor from Intel Xeon ^®^ W-2123 CPU 3.60 GHz, 16GB of 2666MHz DDR4 RAM, 500GB SATA 2.5-inch solid-state drive, and NVIDIA Quadro P620 3GB graphic card.

## Results

The effect of the number of epochs, mini-batch sizing and data augmentation was evaluated on the classification of wrist radiographs in eleven CNN architectures. Table 6 and Table 7 present the aggregated best results for each architecture in prediction accuracy and Cohen’s kappa score respectively

**Table 6.** Results of accuracy for eleven Convolutional Neural Networks used to classify the wrist images in the MURA dataset. The best results for each row are highlighted in *italics* and the overall the best results are highlighted in **bold**.

| No. | CNNs | SGDM | ADAM | Rms Prop | Mean | Epoch | Mini-batch Size |
| --- | --- | --- | --- | --- | --- | --- | --- |
| 1 | GoogleNet | 0.650 | *0.671* | 0.640 | 0.654 | 30 | 64 |
| 2 | VGG-19 | 0.680 | *0.681* | 0.590 | 0.650 | 30 | 64 |
| 3 | AlexNet | 0.674 | *0.690* | 0.657 | 0.674 | 50 | 128 |
| 4 | SqueezeNet | 0.683 | 0.657 | *0.690* | 0.677 | 30 | 64 |
| 5 | ResNet-18 | 0.704 | *0.709* | 0.668 | 0.693 | 30 | 64 |
| 6 | Inception-v3 | *0.710* | 0.689 | 0.707 | 0.702 | 10 | 64 |
| 7 | ResNet-50 | 0.686 | *0.718* | 0.716 | 0.707 | 30 | 64 |
| 8 | VGG-16 | 0.692 | 0.713 | *0.716* | 0.707 | 30 | 128 |
| 9 | ResNet-101 | *0.715* | 0.706 | 0.701 | 0.707 | 30 | 32 |
| 10 | DenseNet-201 | *0.733* | 0.695 | 0.722 | 0.717 | 30 | 32 |
| 11 | Inception- ResNet-v2 | 0.712 | *0.747* | 0.710 | 0.723 | 30 | 32 |
| 12 | ResNet-50 (augmentation) | 0.835 | 0.854 | 0.847 | 0.845 | 30 | 64 |
| 13 | Inception-ResNet-v2 (augmentation) | 0.842 | **0.869** | 0.860 | 0.857 | 30 | 32 |

**Table 7.**
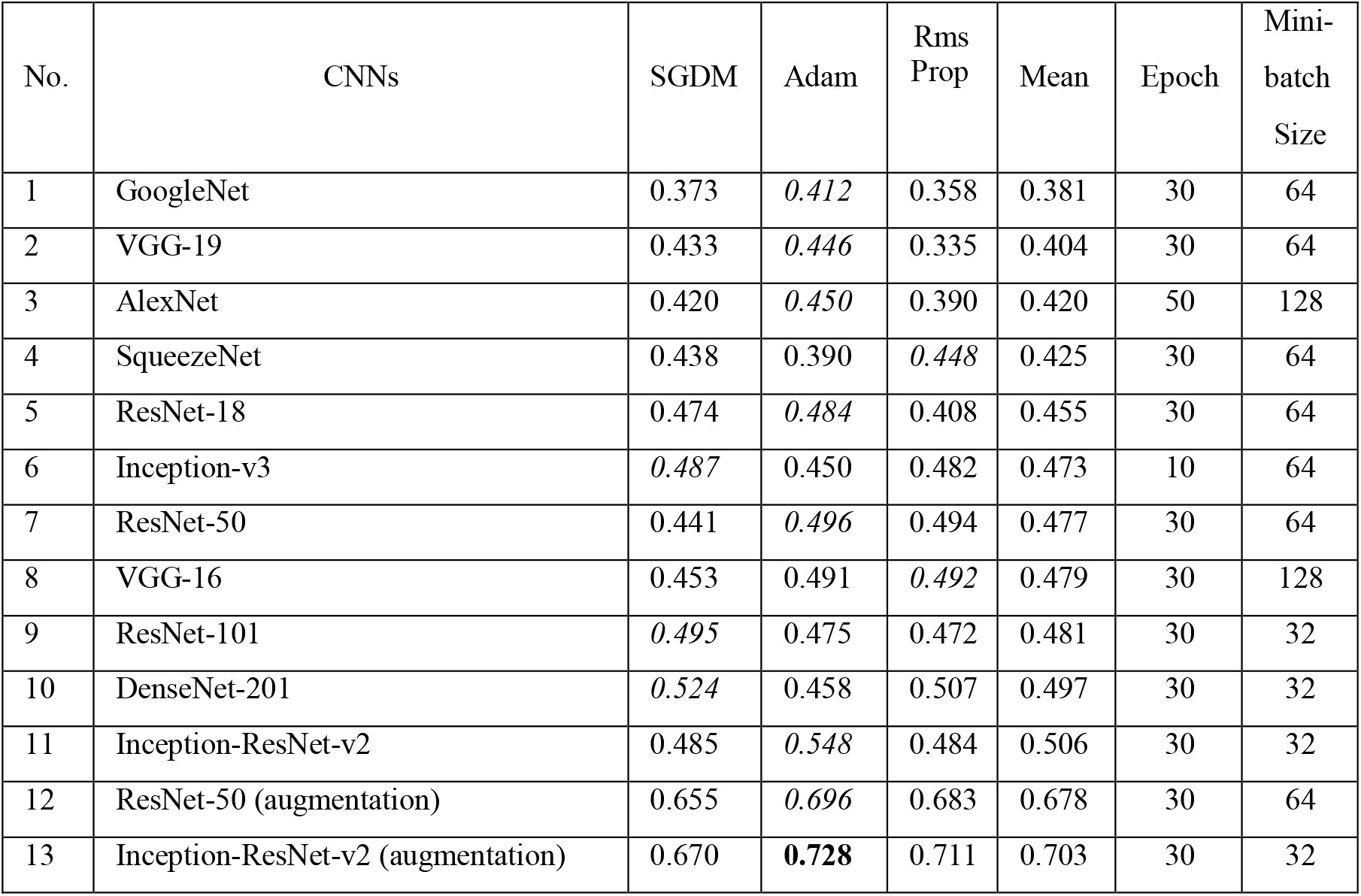
Cohen’s kappa results from eleven Convolutional Neural Networks used to classify the wrist images in the MURA dataset. The best results for each row are highlighted in *italics* and the overall best results are highlighted in **bold**.

| No. | CNNs | SGDM | Adam | Rms Prop | Mean | Epoch | Mini-batch Size |
| --- | --- | --- | --- | --- | --- | --- | --- |
| 1 | GoogleNet | 0.373 | *0.412* | 0.358 | 0.381 | 30 | 64 |
| 2 | VGG-19 | 0.433 | *0.446* | 0.335 | 0.404 | 30 | 64 |
| 3 | AlexNet | 0.420 | *0.450* | 0.390 | 0.420 | 50 | 128 |
| 4 | SqueezeNet | 0.438 | 0.390 | *0.448* | 0.425 | 30 | 64 |
| 5 | ResNet-18 | 0.474 | *0.484* | 0.408 | 0.455 | 30 | 64 |
| 6 | Inception-v3 | *0.487* | 0.450 | 0.482 | 0.473 | 10 | 64 |
| 7 | ResNet-50 | 0.441 | *0.496* | 0.494 | 0.477 | 30 | 64 |
| 8 | VGG-16 | 0.453 | 0.491 | *0.492* | 0.479 | 30 | 128 |
| 9 | ResNet-101 | *0.495* | 0.475 | 0.472 | 0.481 | 30 | 32 |
| 10 | DenseNet-201 | *0.524* | 0.458 | 0.507 | 0.497 | 30 | 32 |
| 11 | Inception-ResNet-v2 | 0.485 | *0.548* | 0.484 | 0.506 | 30 | 32 |
| 12 | ResNet-50 (augmentation) | 0.655 | *0.696* | 0.683 | 0.678 | 30 | 64 |
| 13 | Inception-ResNet-v2 (augmentation) | 0.670 | **0.728** | 0.711 | 0.703 | 30 | 32 |

For the 11 architectures with no data augmentation, Inception-Resnet-v2 performs the best with an accuracy (*Ac* = 0.723) and Cohen’s kappa (*κ* = 0.506). DenseNet-201 fares slightly lower (*Ac* = 0.717, *κ* = 0.497). The lowest results were obtained with GoogleNet (*Ac* = 0.654, *κ* = 0.381). This potentially indicates better feature extraction with deeper network architectures. Fig. 5 and Fig. 6 illustrate some cases of the classification for Lateral and Postero-anterior views of wrist radiographs.

**Figure 5.**
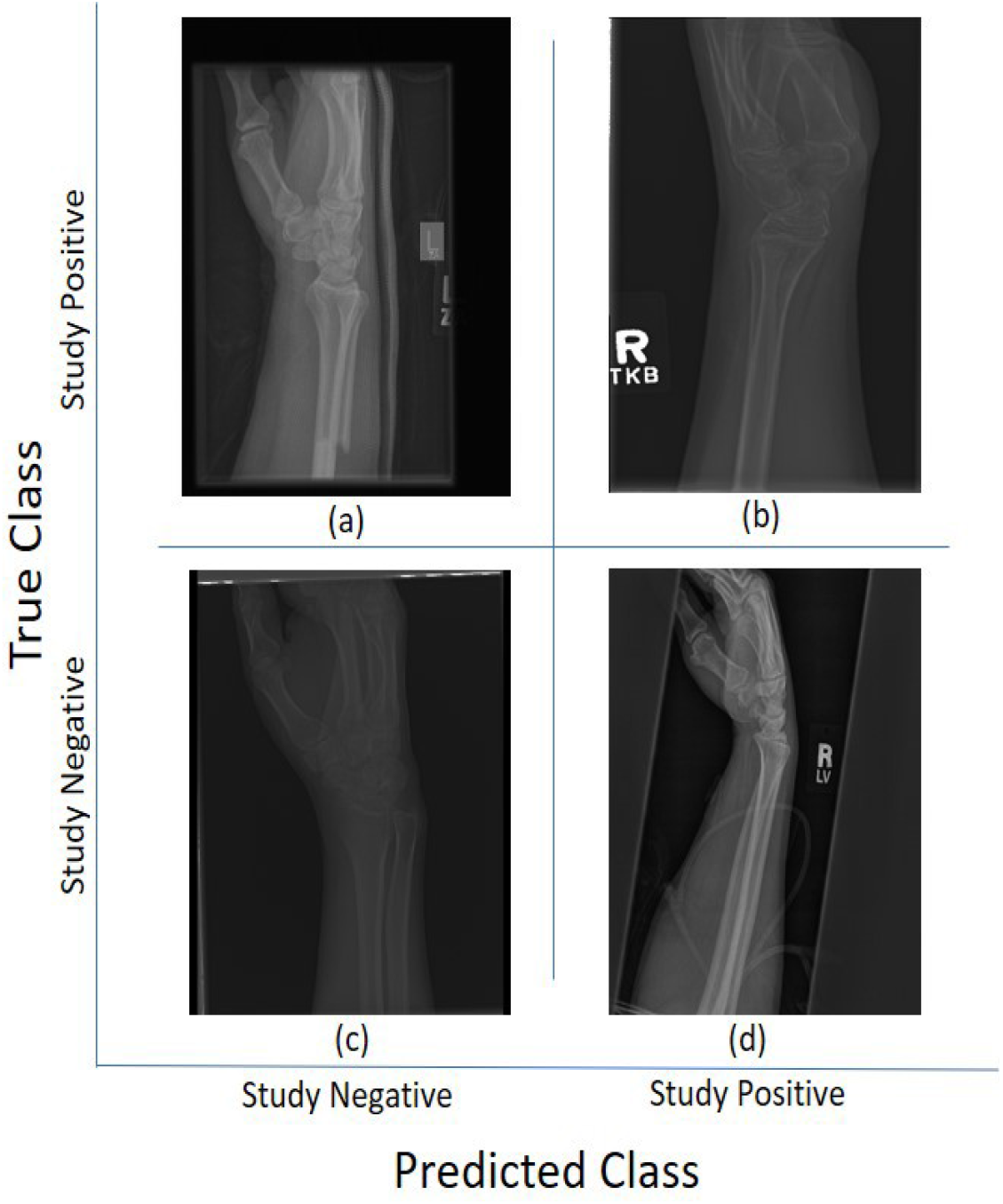
Illustration of classification results for Lateral (LA) views of wrist radiographs. (a) Corresponds to positive (abnormal) diagnosis image but predicted as negative (normal), (b) Abnormal diagnosis and abnormal prediction. (c) Normal diagnosis image and normal prediction. (d) Normal diagnosis and abnormal prediction. Notice that the errors in classification may have been biased by artefact elements on the images.

**Figure 6.**
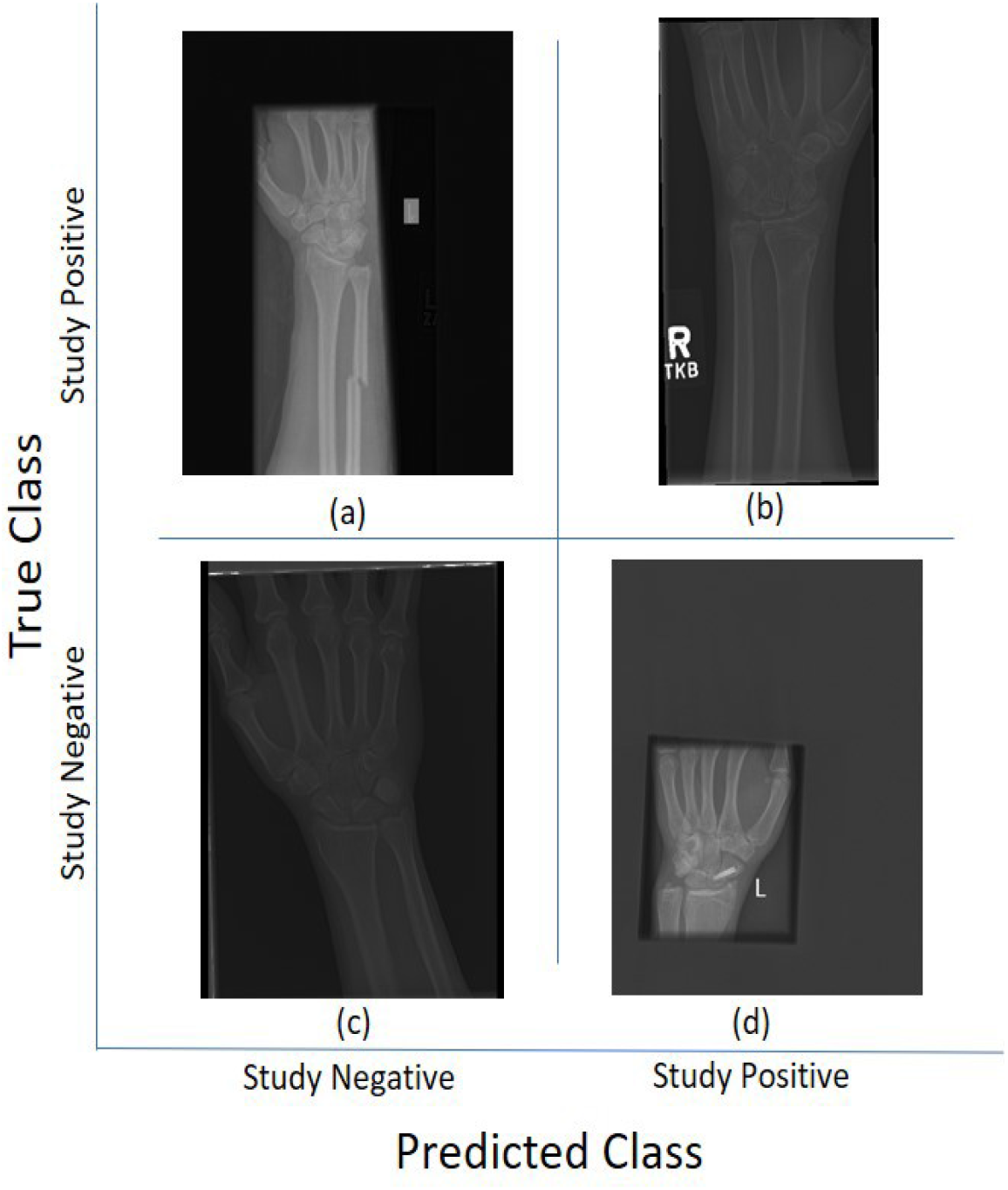
Illustration of classification results for Postero-Anterior (PA) views of wrist radiographs. (a) corresponds to a positive (abnormal) diagnosis image that is predicted as negative (normal); (b) to abnormal diagnosis and abnormal prediction; (c) to normal diagnosis image and normal prediction; and (d) to normal diagnosis and abnormal prediction. Notice again that the errors in classification may have been biased by artefactual elements on the images.

The comparison between ADAM, SGDM, and RMSprop shows no indicative superiority implying that each of these optimisers were capable of achieving the optimal solution. Incremental change to the number of epochs beyond step 30 yields no improvement in accuracy indicating that the models have converged. The choice of the attempted mini-batches shows no difference in results. With data augmentation, the results show significant improvement, e.g., accuracy increases by approximately 19% (up by 0.134) and Cohen’s kappa by 39% (up by 0.197) for the Inception-ResNet-v2 architecture.

Class activation maps were obtained and overlaid on top of the representative images in Figures 1 and 2. The CAMs obtained for ResNet50 are shown in Figures 7 and 9 while those for Inception-ResNet-v2 are shown in Figures 8 and 10. In all cases, the CAMs were capable of indicating the region of attention used in the two architectures applied. This is especially valuable for identifying where the abnormalities are in Figure 9 and 10. While both models indicate similar regions of attention, Inception-ResNet-v2 appears to have smaller attention regions (i.e.,more focused) than those in ResNet50. This may indicate a better extraction of features in the Inception-ResNet-v2 leading to better prediction results.

**Figure 7.**
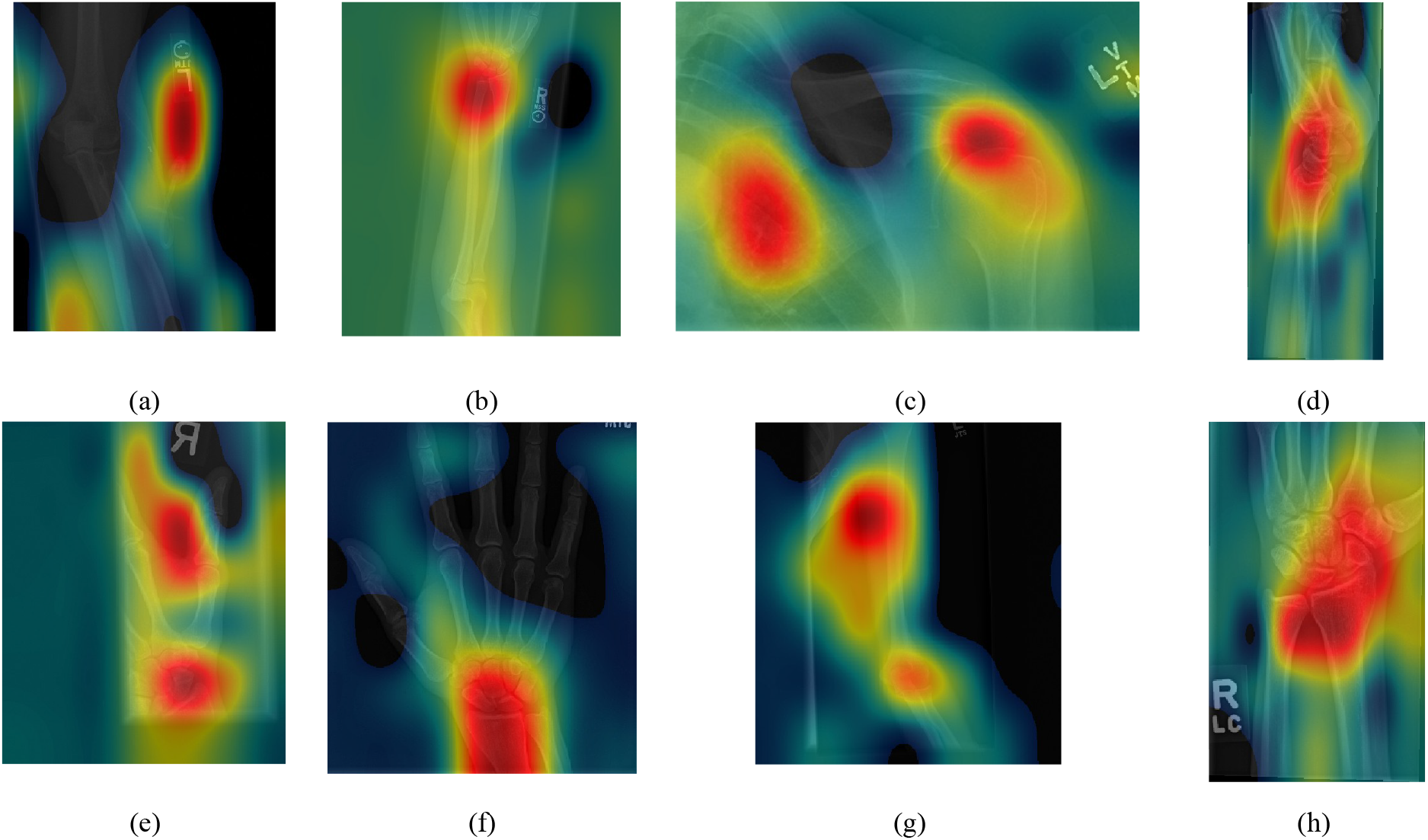
Illustration of Activation Maps overlaid over the eight radiographs without abnormalities of Figure 1 to indicate the regions of the image that activated a **ResNet 50** architecture. (a) Elbow,(b) Forearm, (c) Shoulder, (d) Wrist (lateral view), (d) Lateral view of Wrist, (e) Finger, (f) Hand, (g) Humerus, (h) Wrist. As these cases are positive (no abnormality), the regions of activation are not as critical as those with abnormalities.

**Figure 8.**
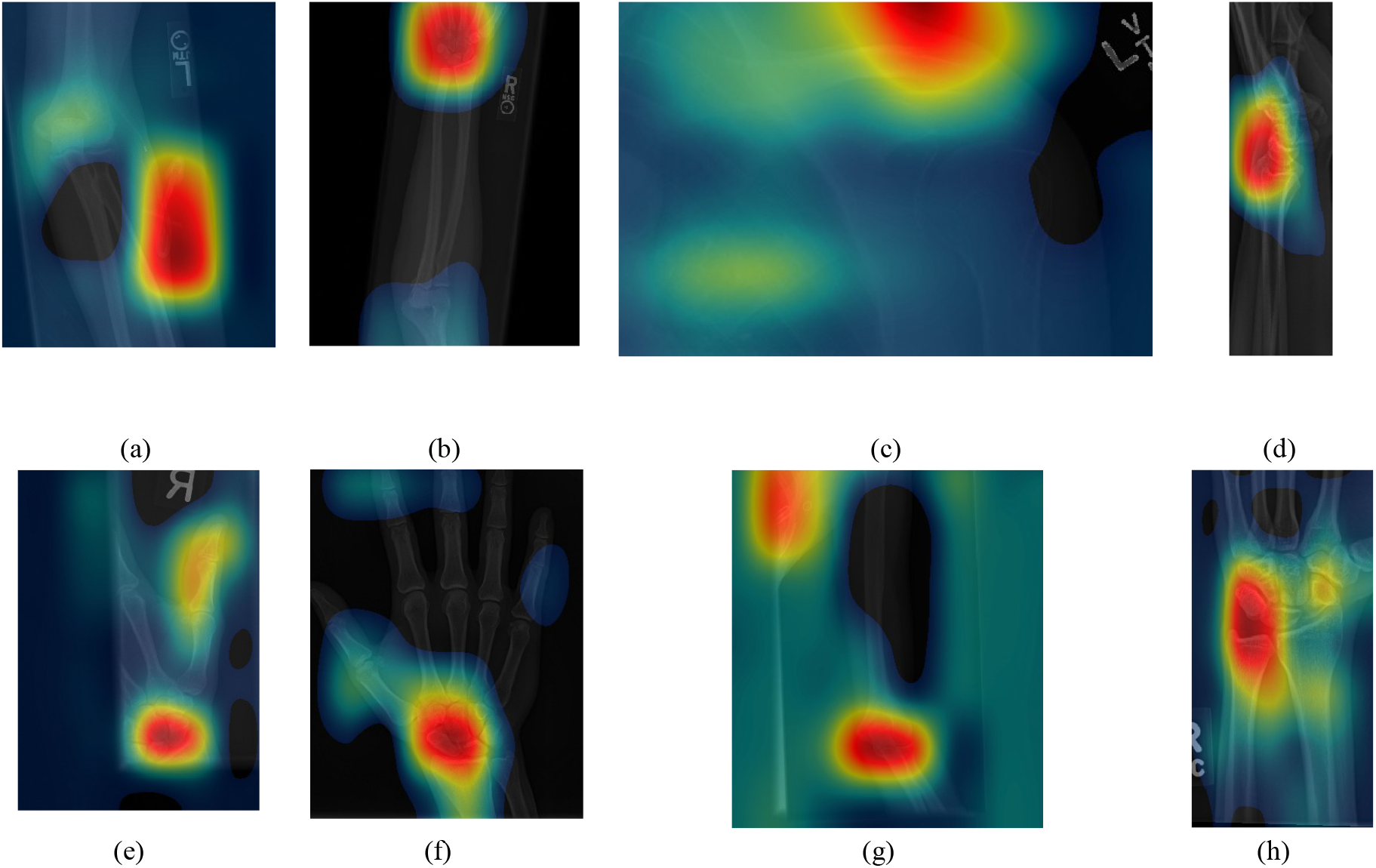
Illustration of Activation Maps overlaid over the eight radiographs without abnormalities of Figure 1 to indicate the regions of the image that activated an **Inception-Resnet-V2** architecture. (a) Elbow, (b) Forearm, (c) Shoulder, (d) Wrist (lateral view), (d) Lateral view of Wrist, (e) Finger, (f) Hand, (g) Humerus, (h) Wrist. It should be noted that the activation regions are more localised than those of the ResNet 50.

**Figure 9.**
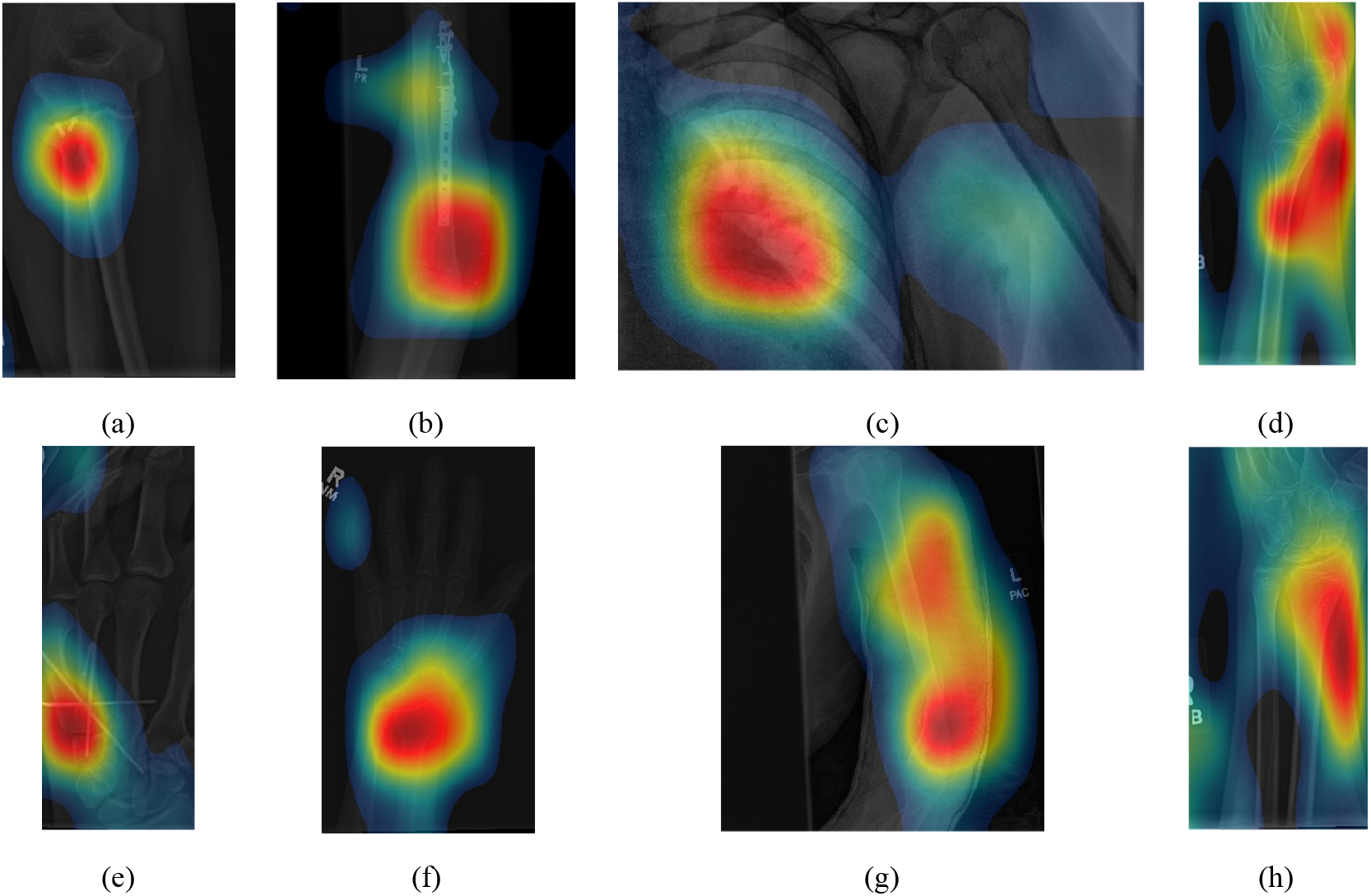
Illustration of Activation Maps overlaid over the eight radiographs with abnormalities of Figure 2 to indicate the regions of the image that activated a **ResNet 50** architecture. (a) Elbow, (b) Forearm, (c) Shoulder, (d) Wrist (lateral view), (d) Lateral view of Wrist, (e) Finger, (f) Hand, (g) Humerus, (h) Wrist. The activation maps illustrate the location of the abnormalities, e.g., (a,e), but appears spread in other cases (b,g) where the abnormality is detected together with a neighbouring region. In other cases (c) the abnormality is not detected.

**Figure 10.**
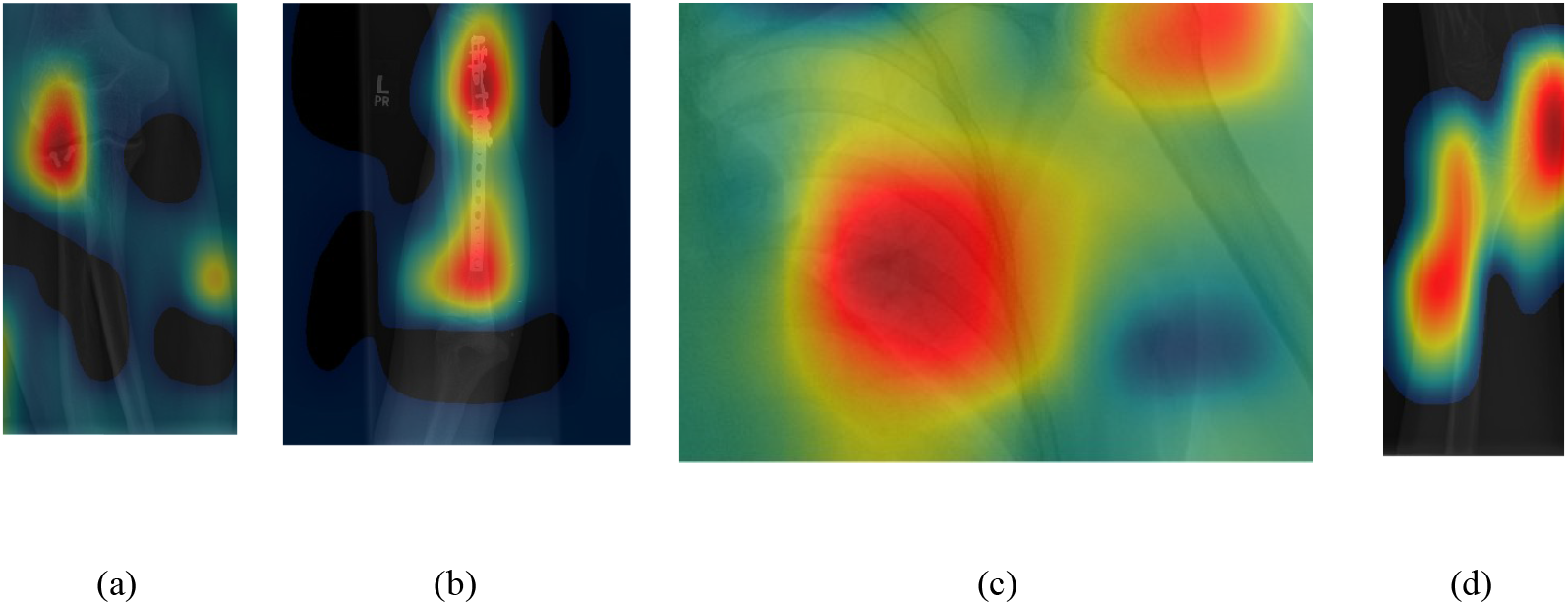
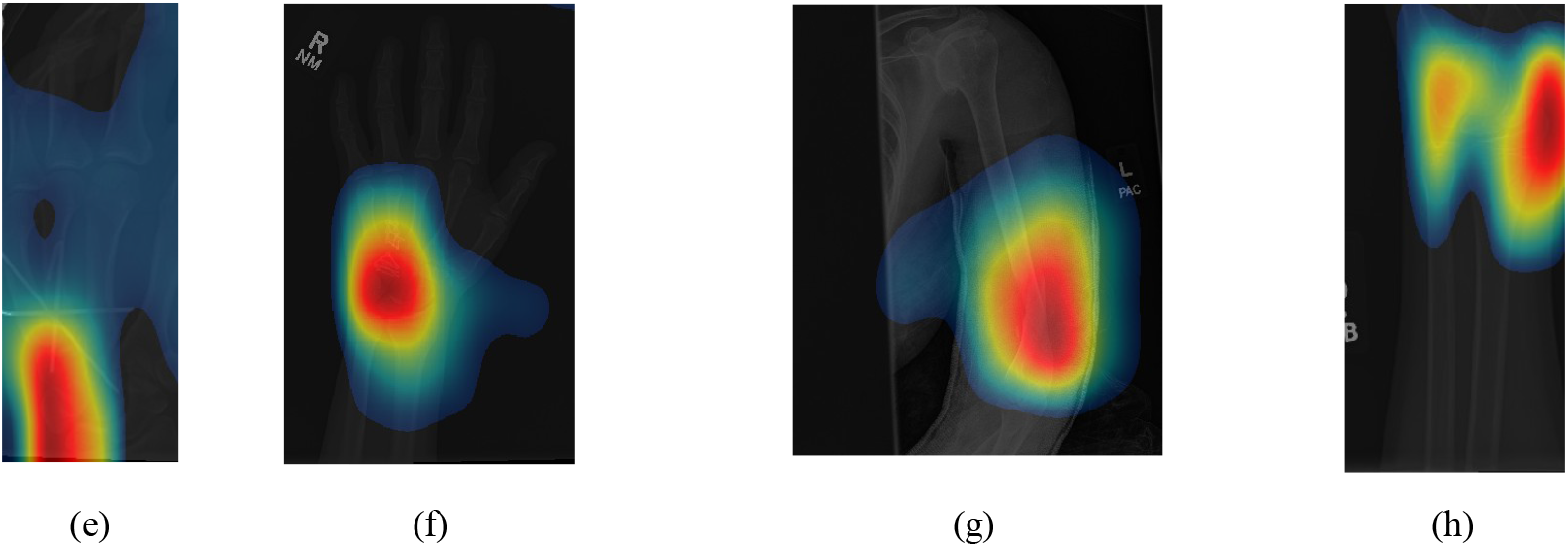
Illustration of Activation Maps overlaid over the eight radiographs with abnormalities of Figure 2 to indicate the regions of the image that activated an **Inception-Resnet-v2** architecture. As for the cases without abnormalities, the activation regions are more located e.g., (g) and in addition, the abnormalities are better located, e.g., (b,c).

## Discussion

In this paper, eleven CNN architectures for the classification of wrist x-rays were compared. Various hyperparameters were attempted during the experiments. It was observed that Inception-Resnet-v2 provided the best results (*Ac* = 0.747, *κ* = 0.548), which were compared with leaders of the MURA challenge which reports 70 entries. The top three places of the leaderboard were *κ* = 0.843, 0.834, 0.833, the lowest score was *κ* = 0.518 and the best performance for a radiologist was *κ* = 0.778. Thus, without data augmentation, the results of all the networks were close to the bottom of the table. Data augmentation significantly improved the results to achieve the 25^*th*^ place of the leaderboard with (*Ac* = 0.869, *κ* = 0.728). Whilst this result was above the average of the table, the positive effect of data augmentation was confirmed to be close to human-level performance.

The CAM provides a channel to interpret how a CNN architecture is trained for feature extraction and the visualisation of the CAMs in the representative images was interesting in several aspects. First, the activated regions in ResNet-50 appeared more broad-brushed than those of the Inception-Resnet-v2. This applied both to the cases without abnormalities (Figures 7 and 8) and those with abnormalities (Figures 9 and 10); Second, the localisation of regions of attention by Inception-Resnet-v2 also appeared more precise than the ResNet-50. This can be appreciated in several cases, for instance the forearm that contains a metallic implant (b) and the humerus with a fracture (g); Third, the activation on the cases without abnormalities provides a consistent focus in areas where abnormalities are expected to appear. This suggests that the network has appropriately learned regions essential to the correct class prediction.

One important point to notice is that all the architectures provided lower results than those at the top of the MURA leaderboard table, even those tested with data augmentation. Whilst in this paper only the wrist subset of the MURA dataset was analysed, it is not considered that these would be more difficult to classify than other anatomical parts. When data augmentation was applied to the input ofthe architectures, the results were significantly better, but still lower than the leaderboard winners. We speculate further steps could improve the performance of CNN-based classification. Specifically:

1. **Data Pre-Processing**: In addition to a grid search of the hyper-parameters, image pre-processing to remove irrelevant features (e.g. text labels) may help the network to target its attention. Appropriate data augmentations (e.g. rotation, reflection, etc) will allow better pattern recognition to be trained and, in turn, provides higher prediction accuracy.
2. **Post Training Evaluation**: Class Activation Map provides an interpretable visualisation for clinicians and radiologists to understand how a prediction was made. It allows the model to be re-trained with additional data to mitigate any model bias and discrepancy. Having a clear association of the key features with the prediction classes [70] will aid in developing a more trustworthy CNN-based classification especially in a clinical setting.
3. **Model Ensemble** [71,72] or combination of the results of different architectures have also shown better results than an individual configuration. This is also observed in the leaderboard for the original MURA competition.
4. **Domain Knowledge**: The knowledge of anatomy (e.g. bone structure in elbow or hands [73]) or the location/orientation of bones [28] can be supplemented in a CNN-based classification to provide further fine tuning in anomaly detection as well as guiding the attention of the network for better results [74].

## Data Availability

The dataset for this study is publicly available by request from Stanford Machine
Learning Group at https://stanfordmlgroup.github.io/competitions/mura/

https://stanfordmlgroup.github.io/competitions/mura/

## Supplementary Materials

The dataset for this study is publicly available by request from Stanford Machine Learning Group at https://stanfordmlgroup.github.io/competitions/mura/

## Abbreviations

The following abbreviations are used in this manuscript:

Ac: Accuracy
A&E: Accidents and Emergency
AI: Artificial Intelligence
CAM: Class Activation Mapping
CNN: Convolutional Neural Network
CT: Computed Tomography
ILSVRC: ImageNet Large Scale Visual Recognition Challenge
MUA: Manipulation under Anaesthesia
MURA: Musculoskeletal Radiographs
ORIF: Open Reduction and Internal Fixation
ReLU: Rectified Linear Unit

